# Utilization of Electronic Health Records for the Assessment of Adiponectin Receptor Autoantibodies during the Progression of Cardio-metabolic Comorbidities

**DOI:** 10.1101/2020.03.09.20033431

**Authors:** Michael J. Pugia, Meeta Pradhan, Rong Qi, Doreen L. Eastes, Anna Geisinger, Bradley J. Mills, Zane Baird, Aruna Wijeratne, Scott M. McAhren, Amber L. Mosley, Anantha Shekhar, Daniel H. Robertson

**Affiliations:** Bioanalytical Research Core, Indiana Biosciences Research Institute, Indianapolis IN; Applied Data Sciences Center, Indiana Biosciences Research Institute, Indianapolis IN; Indiana University School of Medicine, Indianapolis IN; Eli Lilly and Company, Lilly Corporate Center, Indianapolis IN

## Abstract

**BACKGROUND:** Diabetes is a complex, multi-symptomatic disease that drives healthcare costs through its complications as the prevalence of this disease grows rapidly world-wide. Real-world electronic health records (EHRs) coupled with patient biospecimens, biological understanding, and technologies can lead to identification of new diagnostic markers.

**METHODS:** We analyzed the 20-year EHRs of 1862 participants with midpoint samples (10-year) in an observational study of type 2 diabetes and cardiovascular arterial disease (CVAD) conducted by the Fairbanks Institute to test the diagnostic biomarkers. Participants were assigned to four cohorts (healthy, diabetes, CVAD, CVAD+diabetes) based on EHR data analysis. The immunoassay reference range for circulating autoantibodies against the C-terminal fragment of adiponectin receptor 1 (IgG-CTF) was determined and used to predict outcomes post-sample.

**RESULTS:** The IgG-CTF reference range was determined [75–821 ng/mL] and out-of-range values of IgG-CTF values predicted increased likelihood of additional comorbidities and mortality determined from the EHRs 10 years after sample collection. The probability of mortality was lower in patients with elevated IgG-CTF >821 ng/mL [OR 0.49–0.0] and higher in patients with lowered IgG-CTF <75 ng/mL [OR 3.74–9.64]. Although many patients at the time of sample collection had other conditions (hypertension, hyperlipidemia, or elevated uristatin values), only hypertension correlated with increased likelihood of mortality (OR 4.36–5.34).

**CONCLUSIONS:** This study confirms that retrospective analysis of biorepositories coupled with EHRs can provide insight into novel diagnostic markers and the IgG-CTF marker can predict the likelihood of progressing to additional comorbidities or mortality.

## BACKGROUND

Diabetes is an independent risk factor for cardiovascular arterial disease (CVAD), kidney disease, liver disease, Alzheimer’s disease, and many other comorbidities, the costs of which are increasing at >25% per year, with 380 million people likely to be affected by 2025 (1). CVAD has the greatest economic burden, affecting one in four American adults and accounting for 6 million hospitalizations per year as well as nearly 40% of all deaths (∼17 million per year). Patients with both diabetes and CVAD exhibit significantly higher risks for additional complications than those with diabetes alone (2,3).

Insulin resistance and chronic inflammation are strongly associated with the progression of metabolic syndrome and CVAD in diabetes patients (4,5). These are complex, multi-factorial conditions and numerous biomarkers have been proposed, but few have proven effective for patient management (5). An ideal biomarker would provide risk assessment across all individuals and accurately predict progression to various complications. However, current methods are non-specific and cannot predict progression without complex rule-in and rule-out algorithms. Although anti-hyperglycemic medications and standards of care for the management of weight, diet, glycosylated hemoglobin, micro-albuminuria, and albuminuria significantly reduce the risk of CVAD, there remains an urgent need to improve the health economics of diabetes (6-8).

Clinical outcome studies of diabetic complications can be long and expensive, with the death rate at 1–3% per year (1,9). The cost of diagnostic marker assessment is prohibitive for relative risk analysis unless retrospective studies are used to confirm progression outcomes. Additionally, investigational assessments based on cohort comparison often introduce selection bias and cannot justify the verification and validation costs for prospective analysis (10). Retrospective analysis also lacks real-world content because it pre-defines the outcome and cannot adjust for wider results. The use of electronic heath records (EHRs) provides real-world data, offering a promising and more economical method for the assessment of comorbidities, as recently shown for the prediction of chronic kidney disease (11).

Diagnostic accuracy can be improved by the detection of autoantibodies, as shown for autoantibodies against cytokines that predict autoimmune disease and tissue injury caused by autoreactive antibodies and T cells (12,13). Autoantibodies can be quantified using anti-peptide antibodies, as shown for the adiponectin receptor C-terminal fragment (AdipoR1 CTF_344-375_), which circulates freely in the plasma of healthy individuals but not in some diabetes patients (P>0.001) (14). Proteomic analysis of autoantigens is complex and difficult to translate due to the low transient concentrations (<5 ng/mL) and the need to use both stable isotope standards and monoclonal antibodies for capture (15-17). However, once characterized, autoantigens can lead to the identification of new receptor domains, such as the highly conserved AdipoR1 CTF_351-362_ fragment, a strong non-competitive inhibitor of insulin-degrading enzyme (IDE) (18). Autoantibodies can be measured routinely in human samples due to their high non-transient concentrations, including autoantibodies against AdipoR1 CTF_344-375_ (IgG-CTF) with a concentration range of 5–4900 ng/mL (18).

Measuring the diagnostic significance of autoantibodies as a personalized response to disease is difficult given the long time needed for the disease to develop. However, the interaction of AdipoR1 CTF_351-362_ with IDE has diagnostic potential because mechanistic and drug studies have confirmed an impact of IDE on the insulin response in type 2 diabetes and CVAD (19-22). AdipoR1, a G-protein coupled receptor, enhances glucose uptake and fatty acid oxidation in muscle, suppresses glucose output by the liver, and increases insulin sensitivity (23-26). Low adiponectin levels predict a higher risk of type 2 diabetes and CVAD, and the quantification of autoantibodies against AdipoR1 would thus provide a robust and efficient method to determine the diagnostic significance of autoantibodies (27-35).

The Fairbanks Institute for Healthy Communities established a biorepository comprising samples from more than 1900 Indianapolis-area type 2 diabetes and CVAD patients and controls over a 2-year period. Today, this sample bank combined with the patient’s EHRs allows the analysis of novel biomarkers their utility in predicting diabetic complications. We assess the levels of IgG-CTF autoantibodies in the plasma of healthy controls, diabetes and CVAD patients using the Fairbanks biorepository, in combination with real-world outcomes based on EHR data covering the subsequent 10 years, to establish IgG-CTF reference ranges and understand the risk of additional comorbidities.

## METHODS

### Fairbanks Institute Biorepository

The Fairbanks Institute biorepository (NCT01386801, NCT00741416) was created as an extensively-annotated sample repository for hypothesis-driven research that would lead to advancements in the diagnosis, treatment and prevention of diseases affecting the population of Indiana. The study was conducted in accordance with Indiana University’s Internal Review Board (Protocol 1011003179: Multicenter Research Study to Build a Repository that will allow Researchers to Study Chronic Diseases in the Population of Central Indiana). All participants were 18 years or older and gave consent to provide samples.

The participants of this study were originally recruited to the type 2 diabetes cohort, CVAD cohort or healthy controls to a cohort as defined by the criteria listed below during timeframe of sample collection (2007– 2010). Study subjects in the diabetes cohort were recruited based on a EHR-confirmed history of at least one of the following: fasting blood glucose ≥126 mg/dL on two separate occasions; random (non-fasting) blood glucose ≥200 mg/dL on two separate occasions; blood glucose >200 mg/dL at 2 h during a standard oral glucose tolerance test; or hemoglobin A1c (HbA1c) ≥6.5%. Study subjects in the CVAD cohort were recruited based on an EHR-confirmed history of at least one of the following: angioplasty, with or without stent placement; coronary artery bypass graft surgery; diagnostic angiogram; or positive catheterization results showing ≥50% occlusion. Healthy controls for the study were recruited based confirmed history of not having any form of diabetes (as defined above), CVAD, or other risk factors.

Biological specimens were collected from 1966 individuals (n=724 CVAD, n=590 diabetes, n=652 controls) as follows: three 10-mL EDTA tubes for plasma, 14 mL of urine, three 10-mL serum separation tubes (red tops) for serum, and two 3-mL PAXgene tubes for RNA. Specimens were divided into 0.5-mL aliquots and were stored at –80 °C (BioStorage Inc, Indianapolis, IN). Urine specimens were only collected from the diabetes group and half of the control subjects. Of these specimens, 1862 individuals had a complete set of samples with corresponding EHRs that could be used for the biomarker assessment in this study. The patient demographics at the time of sampling is detailed in Table 1.

**Table 1.** Patient demographics from electronic health records at and after sample collection.

|  |  | Cohorts or affected groups* |  |  |  |
| --- | --- | --- | --- | --- | --- |
|  | Total | Control | Diabetes<br>(no CVAD) | CVAD<br>(no diabetes) | CVAD+<br>diabetes |
| **N** | **1862** | **733** | **427** | **344** | **358** |
| **N, Sample + 5 years** |  | 607 | 415 | 340 | 500 |
| Gender, M (%) | 943 (50.6%) | 278 (37.9%) | 210 (49.2%) | 245 (71.2%) | 210 (58.7%) |
| Gender, F (%) | 919 (49.4%) | 455 (62.1%) | 217 (50.8%) | 99 (28.8%) | 148 (41.3%) |
| Age (SD) | 56.1 (9.7) | 54.6 (9.3) | 54.2 (9.3) | 58.5 (9.8) | 59.3 (9.8) |
| Age range | [22 - 83] | [34 - 82] | [36 - 82] | [30 - 83] | [22 - 82] |
| Race, white (%) | 1525 (81.9%) | 551 (75.2%) | 350 (82.0%) | 305 (88.7%) | 319 (89.1%) |
| Race, black (%) | 100 (5.4%) | 35 (4.8%) | 23 (5.4%) | 15 (4.4%) | 27 (7.5%) |
| Race, other/unknown (%) | 237 (12.7%) | 147 (20.1%) | 54 (12.6%) | 24 (7.0%) | 12 (3.4%) |
| # Deaths (%) | 68 (3.6%) | 11 (1.5%) | 8 (1.6%) | 15 (4.4%) | 34 (9.5%) |
| Age at death (SD) | 67.4 (12.7) | 68.7 (13.3) | 74.9 (12.2) | 70.1 (11.1) | 64.2 (12.7) |
| **Medications (filled) †** |  |  |  |  |  |
| Anti-inflammatory | 751 (40.3%) | 251 (34.2%) | 184 (43.1%) | 142 (41.3%) | 174 (48.6%) |
| Anti-hypertensive | 705 (37.9%) | 64 (8.7%) | 243 (56.9%) | 184 (53.5%) | 214 (59.8%) |
| Anti-hyperlipidemic | 510 (27.4%) | 32 (4.4%) | 176 (41.2%) | 142 (41.3%) | 160 (44.7%) |
| Anti-thrombotic | 62 (3.3%) | 12 (1.6%) | 8 (1.9%) | 13 (3.8%) | 29 (8.1%) |
| Glucose lowering | 461 (24.8%) | 0 (0.0%) | 275 (64.4%) | 0 (0.0%) | 186 (52.0%) |
| **Diagnoses ‡** |  |  |  |  |  |
| Type 2 diabetes | 644 (34.6%) | 0 (0.0%) | 301 (70.5%) | 0 (0.0%) | 343 (95.8%) |
| Cardiovascular disease | 702 (37.7%) | 0 (0.0%) | 0 (0.0%) | 344 (100.0%) | 358 (100.0%) |
| Chronic kidney disease | 106 (5.7%) | 3 (0.4%) | 12 (2.8%) | 20 (5.8%) | 71 (19.8%) |
| Liver disease | 60 (3.2%) | 4 (0.5%) | 21 (4.9%) | 7 (2.0%) | 28 (7.8%) |
| Obese | 315 (16.9%) | 30 (4.1%) | 79 (18.5%) | 46 (13.4%) | 160 (44.7%) |
| Hyperlipidemia | 972 (52.2%) | 88 (12.0%) | 239 (56.0%) | 305 (88.7%) | 340 (95.0%) |
| Hypertension | 786 (42.2%) | 40 (5.5%) | 184 (43.1%) | 244 (70.9%) | 318 (88.8%) |
| **Diagnoses ‡, Sample + 5 years** |  |  |  |  |  |
| Type 2 diabetes | 798 (42.9%) | 0 (0.0%) | 323 (77.8%) | 0 (0.0%) | 475 (95.0%) |
| Cardiovascular disease | 840 (45.1%) | 0 (0.0%) | 0 (0.0%) | 340 (100.0%) | 500 (100.0%) |
| Chronic kidney disease | 207 (11.1%) | 3 (0.5%) | 20 (4.8%) | 29 (8.5%) | 155 (31.0%) |
| Liver disease | 106 (5.7%) | 6 (1.0%) | 33 (8.0%) | 14 (4.1%) | 53 (10.6%) |
| Obese | 462 (24.8%) | 38 (6.3%) | 119 (28.7%) | 48 (14.1%) | 257 (51.4%) |
| Hyperlipidemia | 1176 (63.2%) | 120 (19.8%) | 275 (66.3%) | 300 (88.2%) | 481 (96.2%) |
| Hypertension | 1007 (54.1%) | 51 (8.4%) | 229 (55.2%) | 266 (78.2%) | 461 (92.2%) |

\* Comparisons were made between the control (unaffected) population and the groups with type 2 diabetes, cardiovascular disease (CVAD) or both (CVAD+diabetes). The number of patients with the parameter and the percentage of population with the parameter are shown.
† Diabetes medication included glucose lowering drugs and insulin. Anti-inflammatories include aspirin (ASA) and non-steroidal anti-inflammatories (NSAIDs). Lipid-reducing medications include statins. Anti-hypertensives included ACE inhibitors and/or beta blockers
‡ Clinical diagnosis based on ICD9 and ICD10 codes were used for type 2 diabetes, CVAD, chronic kidney disease, and liver disease. A systolic blood pressure >140 mmHg, a diastolic blood pressure > 90 mmHg, and/or the use of anti-hypertensives defines a hypertensive patient. Patients with a value >5 and/or using lipid reducers are defined as hyperlipidemic. Obesity defined as a body mass index (BMI) > 30. The number and percentage for clinical diagnoses of diabetes, CVAD and CVAD+diabetes are shown at time of sample collection and 5 years later.

### Collection of EHRs

When the Fairbanks Institute was launched, the Regenstrief Institute, Inc. (Indianapolis, IN) was responsible for granting access to EHRs for research use, and it maintained the linkage of sample IDs to patient IDs within the Indiana Network of Patient Care (INPC) EHR repository. The Indiana Health Information Exchange is the organization responsible for collecting the EHRs into the INPC database from the participating healthcare organizations within Indiana. For the available samples, all data over 20 years was requested, including demographics, diagnosis codes, medications, clinical laboratory results, and procedures (for n=1907 patients). No *a priori* filtering of the health data was requested. The data were securely delivered to the IBRI as a set of de-identified data extracts according to Health Insurance Portability and Accountability Act safe harbor rules.

### Initial Analysis of EHR Data

EHRs were cleaned and analyzed to understand the patient disease state at sample collection by mapping diagnosis codes (ICD-9/ICD-10) to diseases. Any recorded diagnosis code prior to or within 30 days after sample collection attributed that disease to that individual at the time of sampling. The patients and control diagnosis codes were reassessed 5 years after sample collection (Table 1). The number of medications was computed by mapping the National Drug Code to pharmaceutical class using the National Library of Medicine RxNorm (36). Due to the variability in duration of prescriptions, a prescription was counted if filled 180 days before or after sample collection. Finally, the clinical variables from the EHRs were normalized relative to names and units of measures, outliers were removed, and the values were computed for each patient as the mean of values within 180 days of the sample collection date.

Diagnostic testing performed during patient care after sample collection was obtained from the EHRs and matched with the demographics for (n=1847) patients and controls who remained in the system (Table 2). Because real-world can be incomplete, not all data was available for each diagnostic test or individual. The absence of data does not necessarily mean the absence of the condition, so the data in Table 2 are considered minimum values. Clinical parameters with fewer than 10 patients per group were not analyzed due to lack of data. Means and standard deviations were used from the total sum of results for each patient and across the available test records.

**Table 2.**
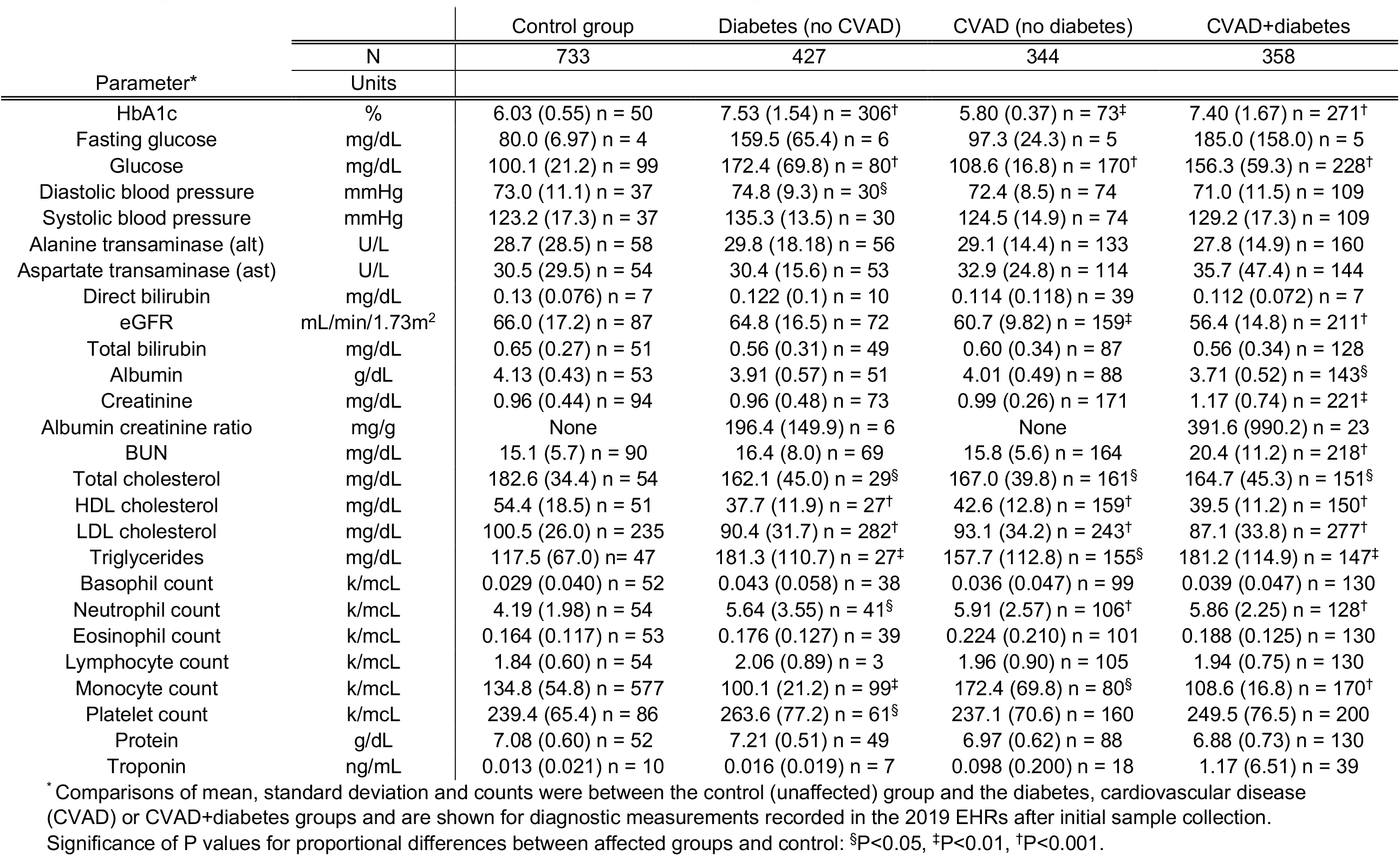
Diagnostic testing data obtained from electronic medical records during patient care.

### Statistical Analysis of EHRs

The affected groups were compared to the control groups for all EHR diagnostic measurements (n=1847) (Table 2). The P value for differences between the affected and control groups was calculated using the normal approximation to the binomial distribution with continuity correction. For quantitative factors (e.g. age and diagnostic tests), the count, mean, standard deviation, minimum and maximum values, and calculated P values were determined for differences between group means assuming a *t*-distribution. Before calculating the *t* value, the group standard deviations were compared using the F distribution. The clinical variables associated with the cohorts defined in Table 1 were analyzed to understand their significance within and across the cohorts. To understand the significance of the variable with respect to each cohort, between cohorts and across cohorts, the data were processed by univariate and bivariate analysis and ANOVA. Each variable in was analyzed for significance across cohorts using an independent samples *t*-test. Table 2 shows the significant P values for each variable. Differences between the affected and control groups were deemed statistically significant if the corresponding P values were less than 0.001 (99.9%), 0.01 (99%) or 0.05 (95%).

### Analysis of EHR Outcomes

The EHRs of total patients (n=1847) and cohorts were assessed for disease indications and the presence of hypertension (HT) and/or hyperlipidemia (HLD) prior to sample collection. The post-sample development of comorbidities was assessed by EHR data analysis to confirm the development of diabetes (n=174), CVAD (n=140), chronic kidney disease (CKD, n=136), or liver disease (LD, n=53). The development of multiple comorbidities such as CVAD+diabetes (n=39), CKD+diabetes (n=32), LD+diabetes (n=10), and CVAD+CKD (n=29) was also assessed. The disease progression from initial sample collection to final outcome based on available EHR data is depicted in the Sankey flow diagram (Figure 1). Additional phenotypes combining more than two comorbidities represented less than 0.7% of the patients and their significance could not be assessed.

**Figure 1.**
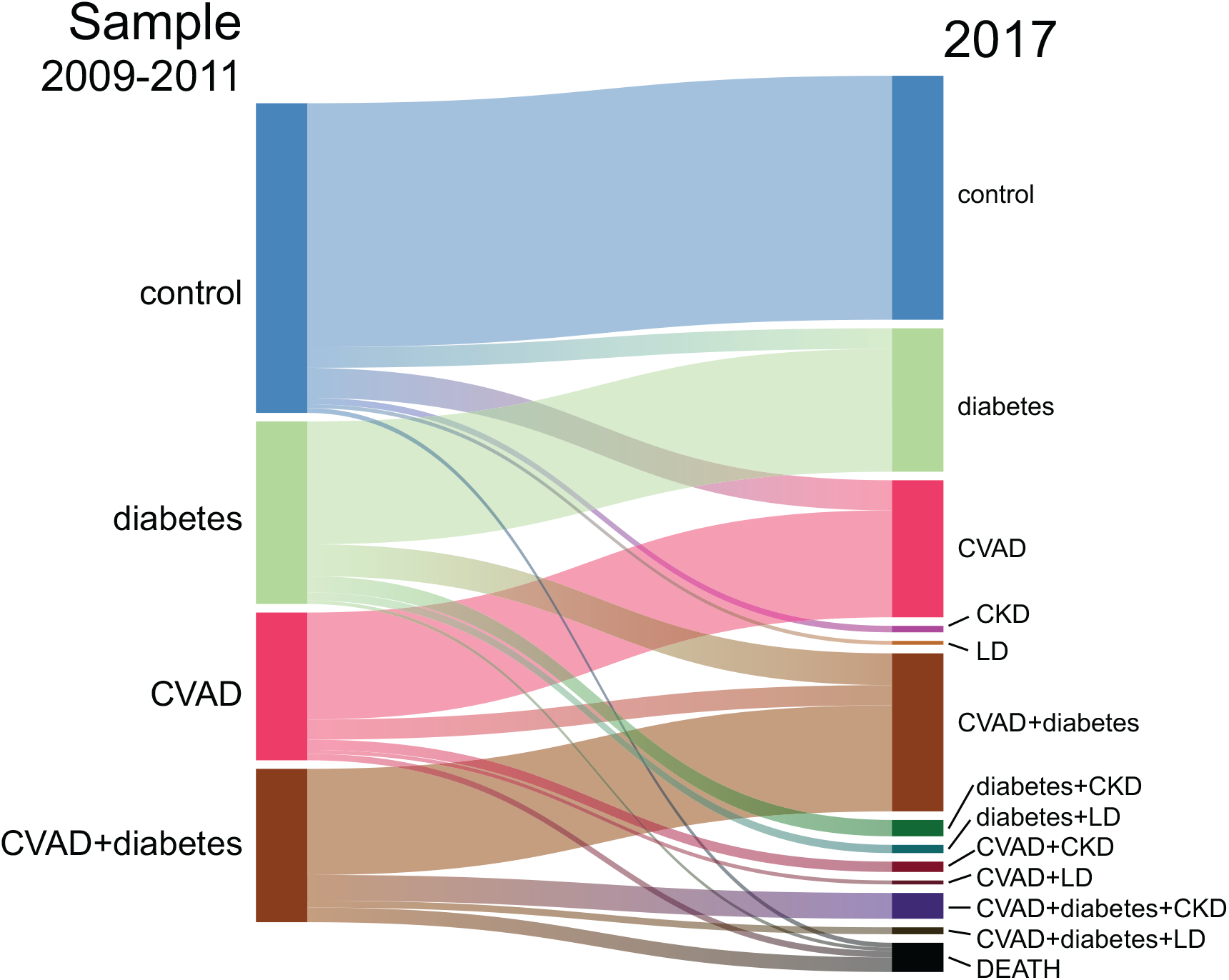
Sankey flow diagram showing the transition from initial disease state at sample collection to final outcome based on EHR data for the 1847 patients. The bars represent the number of individuals within that disease state and the width of the connections represents that number of the individuals transitioning to the new final state after sample collection. The number of individuals with hypertension alone, hyperlipidemia alone, or both, was also computed for these cohorts at sample collection but the data are not shown. These numbers are for the control (29,78,39), diabetes (28, 82, 182), CVAD (16, 77, 242) and CVAD+diabetes (14, 37, 315) cohorts, respectively.

### New Marker Measurements

The presence of any infection or inflammation (Table 3) was determined for all available urine samples (n=663) collected from the T2D cohort by measuring uristatin by competitive enzyme-linked immunosorbent assay (ELISA) as previously described (37). Urine samples (∼3 mL) were thawed to room temperature, 10 µL was transferred to duplicate lanes of a polypropylene sample plate. Uristatin antibody (ATCC 421-3G5.4C5.3B6) conjugated to alkaline phosphatase (ALP) was mixed with 300 mL Super blocker (final concentration = 0.084 µg/L) and 990 µL was mixed with the sample in each well. The sample plate was then sealed with a tight foil cover and incubated at 35 °C for 60 min in a heat block. The plate was stored at 4 °C for testing within 24 h or at –80 °C for future testing (up to 5 years).

**Table 3.**
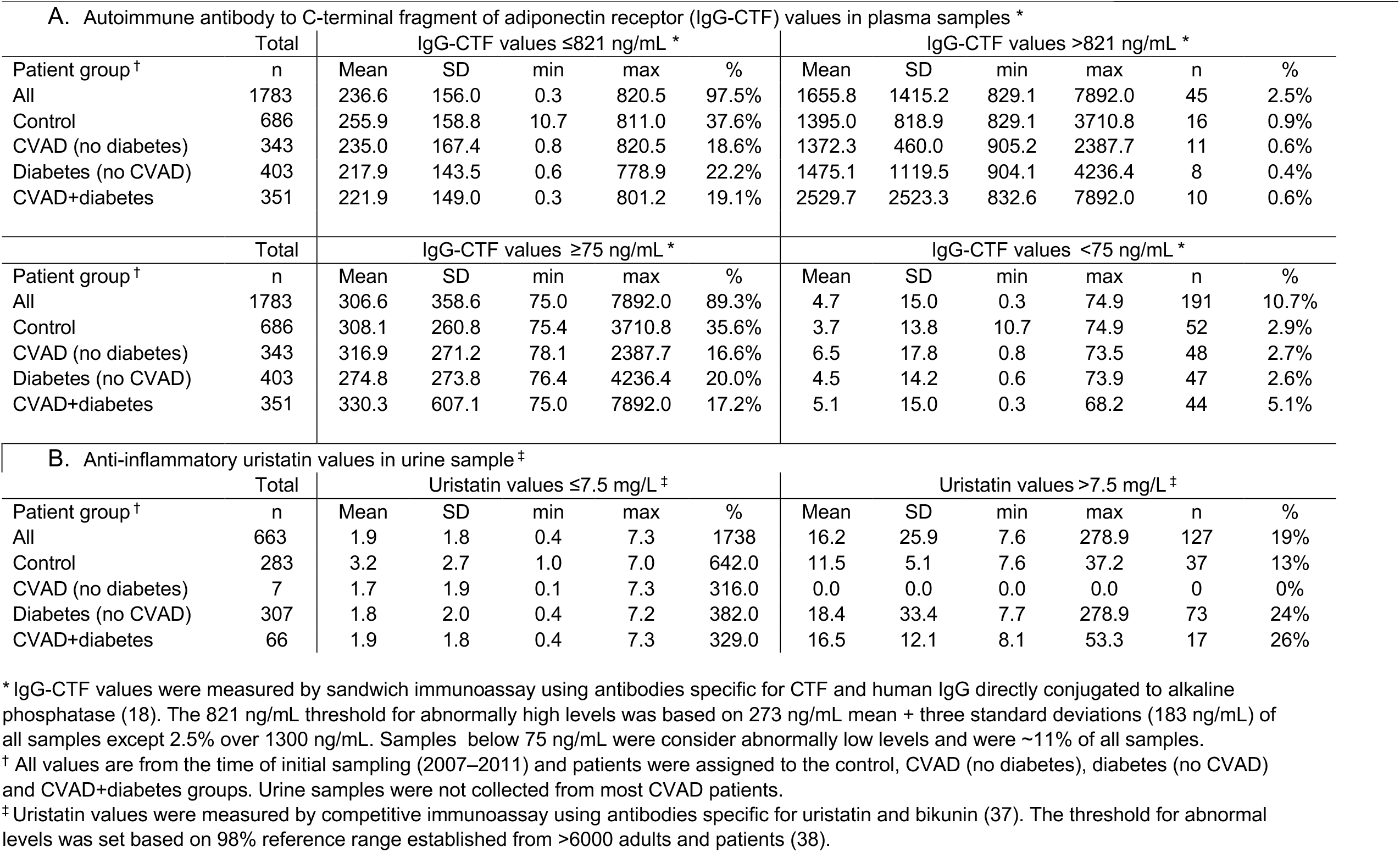
Comparison of Ig-CTF and uristatin reference ranges for values from biorepository samples.

Available plasma specimens (n=1783) were measured for IgG-CTF using a previously described sandwich ELISA based on a monoclonal antibody specific for AdipoR CTF (ATCC 444-1D12.1H7) and a human IgG-specific antibody conjugated to ALP (14) (Table 3). Plasma (∼0.5 mL) was thawed to room temperature, and 10 µL was diluted with 480 µL of stabilization buffer (PBS supplemented with 1% BSA, 0.1 M citrate and 0.01 M EDTA, pH 6.4). Diluted specimens were tested as above or stored at –80 °C.

Proteomics analysis confirming the autoantigen presence in the samples was performed using an anti-AdipoR CTF antibody (444 clone, 4.3 mg/mL) directly conjugated to biotin to extract the IgG-CTF from plasma samples, and the free CTF was then released using Tris base (pH ≥8.3). The free CTF was directly measured against synthetic peptide standards (Celtek Biosciences, Franklin, TN) to simultaneously quantify the AdipoR1, AdipoR2 CTF_343-375_ (32-mer peptides), and AdipoR1 CTF_344-375_ (31-mer peptide) as previously described (14,18). Western blot analysis of plasma samples confirmed the presence of gamma heavy chains (50–55 kDa) and kappa light chains (26–28 kDa) for the bound form of IgG-CTF, as previously reported (14).

### Comorbidity Statistical Analysis

Odds ratios were calculated to compare patients with new disease from those with unchanged clinical profiles when abnormal biomarkers, HT or HLD were present. The morbidity for all cases was determined (n=68) and used to calculate survival odds ratios. Patients developing diabetes, CVAD, CKD or LD at any time after sample collection were considered as the combined group of patients who progressed to additional comorbidities. Patient deaths were assessed across all four patient groups for sample sizes above n=10. The significance of odds ratios was estimated using the variance from multiple sample tests from the same patient during the course of at least 3 days for both uristatin and IgG-CTF. Uristatin variance at ∼7.5 mg/L was ±1.5 mg/L, and the IgG-CTF variance at ∼75 and ∼800 ng/mL was ±35 ng/mL and ±80 ng/mL, respectively. The standard error for the HT and HLD odds ratios was based on the sample size.

## RESULTS

### Participant Characteristics

Patients were equally represented in terms of gender (∼50%) and initial disease groupings (∼20% diabetes, CVAD and CVAD+diabetes) (Table 1). Age (30s to 80s) and race (80% white) were also consistent across the groups. Medications were consistent with expected standards of care. The proportion of patients with diabetes, CVAD and CVAD+diabetes increased significantly in the 5 years after sample collection.

### Laboratory Measurements

The types of diagnostic tests used in this study agreed with the standards of care that would be expected for patients with diabetes and CVAD over this study period. Most patients had multiple tests for key monitors such as glucose, complete blood cell count, lipid panels, and kidney function. The additional measurements indicated the correlation between parameters and conditions are shown in Table 2. Fasting blood glucose levels and HbA1c were elevated for diabetes but not CVAD patients. The spot glucose ranges were significantly elevated for diabetes and only slightly elevated for CVAD patients. The kidney function results based on estimated glomerular filtration rate (eGFR) values were significantly worse for CVAD patients but not diabetes patients. Only the CVAD and diabetes groups had higher mean blood urea nitrogen (BUN) values, lower serum creatine, and lower plasma albumin as expected given the worse renal conditions. Micro-albuminuria testing (albumin/creatine ratio) was measured too rarely to be significantly assessed in the EHR data.

The mean neutrophil counts were significantly elevated in both diabetes and CVAD patients. Monocyte counts were lower in diabetes and higher in CVAD patients. Total lipids, LDL, and HDL were significantly lower in diabetes and CVAD patients compared to controls (patients were generally on lipid-lowering medications). Blood pressure ranges showed no significant differences. Triglyceride levels were highest in diabetes patients followed by CVAD patients and were consistently higher than the control group.

### New Biomarker Testing

Uristatin immunoassay values ranged from 0.1 to 278.9 mg/L, with 81% of the patients within the normal range <7.5 mg/mL at sample collection (Table 3). The 98% reference range of <7.5 mg/mL for normal results was previously determined for 6292 patients lacking infection, inflammation or kidney disease (37-40). In these samples, 19% (n=127) contained uristatin ≥7.5 mg/L. The mean values did not differ significantly between the control, diabetes, and CVAD+diabetes cohorts. Elevated uristatin values (≥7.5 mg/L) were more frequently observed in the disease groups (e.g. diabetes = 24% and CVAD+diabetes = 26%) versus the controls (13%). The observed reference range of IgG-CTF was 75–821 ng/mL (mean 237 ng/mL, standard deviation 156) with 2.5% of participants exhibiting elevated autoantibodies (821–7892 ng/mL) and 11% lacking autoantibodies (>75 ng/mL). No significant differences were observed between control, diabetes, CVAD, and CVAD+diabetes groups when autoantibodies were absent (all groups, 2.9– 5.1%) or present at elevated levels (all groups, 0.4–0.9%) (Table 3).

### Diagnostic Phenotyping and Impact

The highest mortality rates (9.5% over 10 years) were observed for diabetes patients who progressed to CVAD followed by CVAD-only patients (4.4%) while the mortality rates for diabetes patients who developed no comorbidities (1.9%) and controls (1.5%) were significantly lower over the same 10-year period (Table 4). For the control cohort, the most likely new comorbidities were CVAD (9.7%), diabetes (6.7%), CKD (2.1%) and LD (1.2%). For the CVAD cohort, the most likely new comorbidities were diabetes (13.7%), CKD (7.0%) and LD (2.6%). For the diabetes cohort, the most likely comorbidities were CVAD (17.4%), CKD (8.9%) and LD (4.5%). For the CVAD+diabetes cohort, the most likely new comorbidities were CKD (16.8%) and LD (4.5%). Patients with HT were more likely to develop comorbidities (odds ratio 4.36–5.34) then patients positive for HLD and uristatin who no significant likelihood to progress (Table 4). Patients lacking IgG-CTF were more likely to progress to comorbidities (odds ratio 3.74–9.64) whereas patients with elevated IgG-CTF were significantly less likely to progress (odds ratio 0.49–0.0) (Table 4). Finally, patients lacking IgG-CTF were the only ones with any significant likelihood to progress to diabetes and CVAD (odds ratio 2.06–2.97).

**Table 4.**
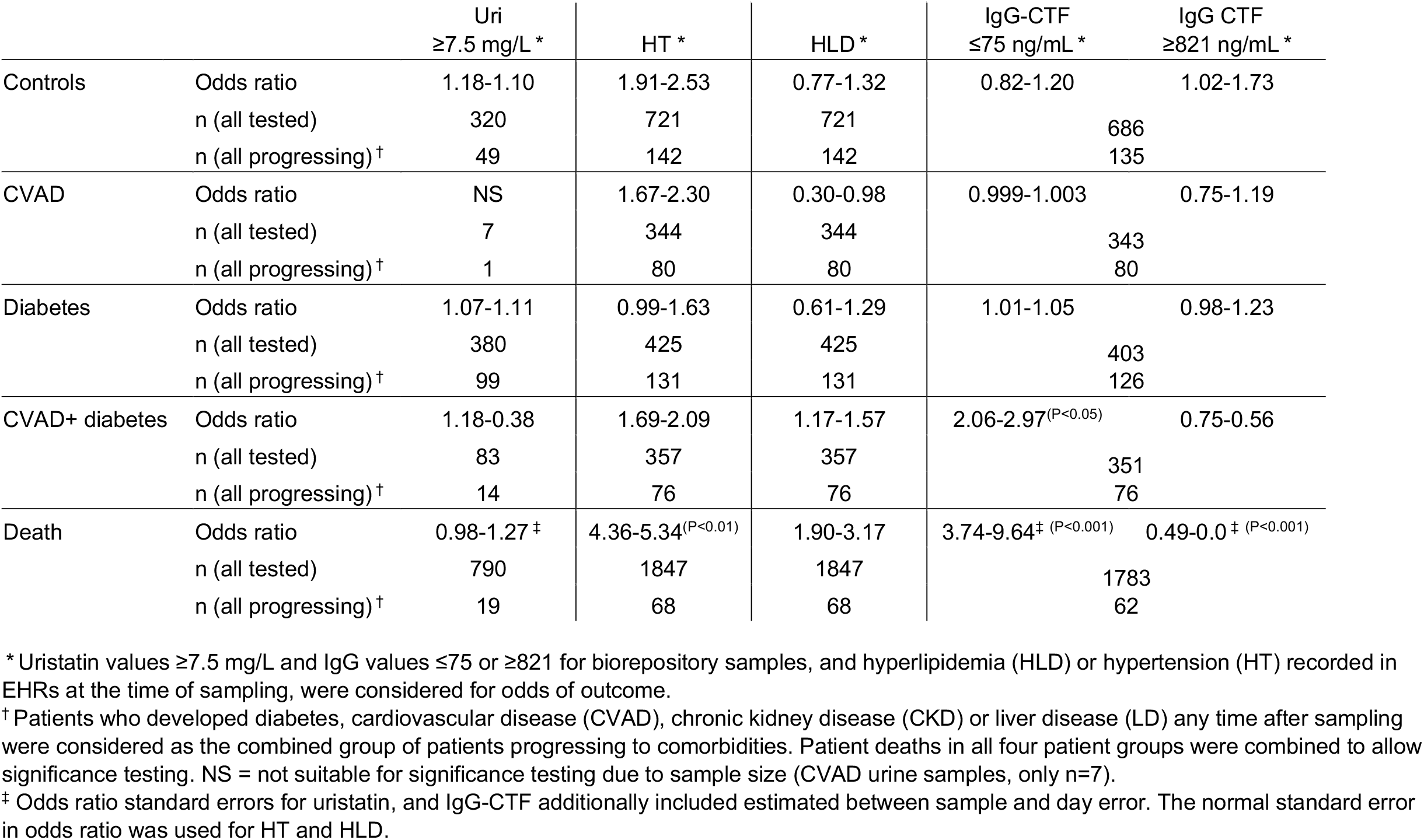
Odds ratios for patient progression to co-morbidities for patients with high blood pressure, lipedema and biomarkers outside of reference ranges.

## CONCLUSIONS

Autoantibody discoveries offer a new generation of potential biomarkers to characterize a wider range of post-translational modifications caused by stress on cells and tissues prior to the development of a diagnosed autoimmune pathology. These methods allow direct observations from human specimens that can uncover unique fragments with biological responses. The discovery of the adiponectin receptor binding to IDE results directly from the proteomic analysis of the AdipoR1 CTF (14). Circulating autoantibodies recognizing this fragment (IgG-CTF) appear in most human and animal blood and were measurable by ELISA in all patients included in this study (18). No significant differences in IgG-CTF values in the diabetes or CVAD disease groups were found compared to healthy controls, which is in agreement with earlier results (18).

The absence of IgG-CTF autoantibodies did correlate significantly with accelerated progression to diabetes and CVAD and a higher risk of mortality. In contrast, patients with elevated levels of IgG-CTF had no deaths and a lower risk of progression to diabetes and CVAD. These findings suggest that IgG-CTF autoantibodies have a neutralizing function, potentially eliminating the ability of CTF peptides to inhibit IDE in the blood. Verification of this new autoantigen in human blood is difficult to translate because the CTF binds other molecules and is only transiently present at very low concentrations, making reproducible measurements difficult to achieve in patient samples (18). However, the measurement of autoantibodies recognizing this proteolytic fragment could potentially indicate receptor turnover due to disease stress early in a patient’s clinical course.

Autoantibody responses can be measured in patients over longer periods of time without seeing significant change. Previous studies showed no variation in the levels of IgG-CTF over 90 days in samples from patients without significant weight change (18). Further work is necessary to understand the development and loss of these autoantibodies during disease. In this study, age, sex, and race did not significantly affect IgG-CTF values in patients and it also agrees with previous findings that autoantibodies in diabetes and control subjects did not correlate with diabetes progression as measured by impaired fasting glucose or impaired glucose tolerance (18). Although IgG-CTF values do not change following glucose administration (GGTT), the free CTF autoantigen does change during the subsequent 120 min in animal models suggesting a biological role (18).

Hypertension was the only other predictor of progression in the EHR data. The levels of uristatin that is formed as part of the anti-inflammatory response (38) did not predict disease progression. Uristatin immunoassay values are <7.5 mg/mL in ∼98% of patients in the absence of infection, inflammation, or kidney disease (38-40), however, we found that significant infection or inflammation was common in diabetes patients (22%) but unable to be assessed in the CVAD cohort due to absence of samples. This lack of correlation between inflammation and the development of comorbidities is unsurprising because a single incidence of inflammation may be quickly resolved is not expected to affect the long-term outcomes.

This study confirms that retrospective analysis of biorepositories using EHR-based outcomes can provide insight into the role of autoantibodies to proteolytic fragments of cell receptors. Autoantibodies immunoassays provide stable measurements and avoid the problems of measuring transient autoantigens. A highly multiplex panel of antibodies covering a wide range of receptor fragments could potentially detect higher receptor turnover in tissues as a more comprehensive multi-factorial receptor response during disease. Genetic blood assays often cannot resolve signals from the post-translational fragmentation of receptors. Here, autoimmune assays may help to resolve phenotypic expression profiles in tissues that cannot be investigated by blood gene testing

## Data Availability

Data is not publically available, but can be obtained under appropriate research agreement and data use agreement.

## Acknowledgments

The authors acknowledge the Regenstrief Institute, Inc. and the Indiana Clinical Translational Sciences Institute for access to the de-identified patient records corresponding to the collected biological specimens. The authors acknowledge BioCrossroads, Troy Hege, and Natalie Stull for the stewardship of the biorepository and transfer for testing. Additionally, we thank our undergraduate intern from IUPUI, Mona Bhattrai, who performed the cell culture for materials for research-use antibodies.

## Author Contributions

M.J.P. contributed to the study design, new marker testing, data analysis, discussion, the acquisition of study support, manuscript writing, editing and review, and the creation of study protocols. D.R. contributed to the study concept, the acquisition of study support, the design, data analytics, the security of the medical records, and manuscript writing and review. M.P., R.Q., B.M., and S.M. recovered and verified the clinical and diagnostic data from the EHRs, performed data analysis, and reviewed and edited the manuscript. D.E. and A.V. collected ELISA data and reviewed the manuscript. Z.B., A.W. and A.M. performed isolation, proteomics, and reviewed and edited the manuscript. A.S. was the Principal Investigator of the Multicenter Research Study to Build a Repository to Study Chronic Diseases in Indiana (IHS) that built the repository. D.R. and M.J.P. are the guarantors of this work and, as such, had full access to all the data in the study and take responsibility for the integrity of the data and the accuracy of the data analysis.

